# Undiagnosed cognitive impairment and willingness to seek help: Community-representative study from Singapore

**DOI:** 10.64898/2026.01.16.26344274

**Authors:** Tau Ming Liew, King Fan Yip, Kaavya Narasimhalu, Simon Kang Seng Ting, Weishan Li, Sze Yan Tay, Way Inn Koay

## Abstract

This study challenges the assumption that undiagnosed cognitive impairment (CI) is driven primarily by patient-level barriers like poor awareness. In a population-weighted cohort of 1,856 older Singaporeans, CI prevalence was 24.7% (95%CI 18.8–31.8); yet the undiagnosed rate was high (81.4%, 95%CI 65.6–90.9), especially for mild CI (97.9%, 95%CI 94.1–99.3). This diagnostic gap persisted despite high symptom awareness (81.3%, 95%CI 63.6–91.5) and help-seeking intent (63.3%, 95%CI 47.5–76.7), with informants becoming key as CI worsened. Findings suggest successful public health campaigns have shifted the bottleneck from community awareness to healthcare system capacity, creating an opportunity for a policy shift to meet rising demand for diagnosis—by empowering primary care with efficient case-finding tools, formalizing integrated diagnostic pathways, and establishing channels for family informants’ involvement. From these findings, we conceptualized a ‘paradox of success’ model, providing a framework for other health systems to adapt policy as public engagement grows.

## INTRODUCTION

Undiagnosed cognitive impairment (CI) is a global challenge,^1^ with 60–90% of affected individuals never receiving a formal diagnosis.^2,3^ Those who remain undiagnosed miss out on timely clinical care—including management of reversible causes, prescription of cognitive enhancers, behavioral management, caregiver support, and advanced care planning^4–7^—which can impact well-being^8,9^ and increase the risk of premature nursing home placement.^10^ Furthermore, undiagnosed individuals often do not receive adequate support to manage and coordinate care for their chronic diseases,^11,12^ resulting in suboptimal disease control, inappropriate healthcare utilization, and higher healthcare costs.^13,14^ In recent years, the importance of timely diagnosis is further accentuated by emerging evidence for early interventions, such as risk factor modification^15,16^ and anti-amyloid monoclonal antibodies.^17,18^

The reasons for undiagnosed CI are complex, but explanations often focused on patient- and family-level barriers.^19^ The prevailing narrative suggests that individuals either lack awareness of their deficits (anosognosia), attribute symptoms to “normal aging”, or avoid diagnosis due to denial, fear, or stigma.^19^ However, this patient-centric perspective is based on an incomplete evidence base. Most research on help-seeking behavior utilized self-selecting, clinic-based samples^19^—that is, individuals who have already successfully navigated the path to diagnosis—and may not reflect the wider undiagnosed population. Furthermore, these studies are mostly from Western populations,^19^ limiting their relevance to Asian contexts where cultural factors (e.g. views on aging and family responsibility) can shape help-seeking.^19,20^ Crucially, few community-based studies have simultaneously assessed the rates of undiagnosed CI, symptom awareness, and willingness to seek help, making it unclear whether the diagnostic gap is driven by patient inaction^19^ or by systemic barriers within the healthcare.^21,22^

To address these gaps, we analyzed data from a large, prospectively recruited cohort of community-dwelling older adults in Singapore, a multi-ethnic and rapidly aging Asian nation. This sample was then re-weighted to be nationally representative, allowing us to pursue two aims:

1) To establish the population-representative prevalence of previously undiagnosed CI (mild cognitive impairment or dementia); and
2) Most centrally, to determine the prevalence of symptom awareness and help-seeking intentions.

By comparing the high proportion of undiagnosed cases with actual help-seeking intent, this study directly tests whether the diagnostic gap stems from patient inaction,^19^ or instead reflects systemic barriers in healthcare^21,22^—a distinction with critical implications for public health policy and practice.

## METHOD

### Study population

This study involved 1,856 community-representative older adults in Singapore, drawn from Project PENSIEVE^23^—a nationally-funded initiative to improve the undiagnosed rates of CI in Singapore.

Participant were prospectively recruited from community settings in Singapore between March 2022 and September 2024. Recruitment sources included 14 community roadshows by the study team, clients of community partners, home visits by community volunteers, media publicity (radio, online articles, and posters), and word-of-mouth referrals from participants who had completed research assessments. Inclusion criteria were: (1) aged ≥65 years; (2) residing in the community (not nursing homes); (3) had at least one of the three chronic diseases: diabetes mellitus, hypertension, or hyperlipidemia; (4) able to follow simple instructions in English or Mandarin; and (5) had a knowledgeable informant (e.g. family member or friend). The sole exclusion criterion was severe visual impairment that preclude neuropsychological testing, applied conservatively to ensure generalizability—participants were included as long as they could see pictures on a piece of paper held in front of them. No participants were excluded for missing data due to mandatory data fields and routine audits.

Ethical approval was granted by the SingHealth Centralized Institutional Review Board (reference: 2021/2590). Informed consent was obtained from all participants or their legal proxies.

### Measures

All participants received comprehensive assessments, which included semi-structured interviews with participants and their informants, detailed neuropsychological testing, and behavioral observations.

Demographic and medical information (e.g. age, education, sex, ethnicity, living arrangements, health conditions) was obtained from participants and informants. Participants’ weight and height were measured to calculate body mass index. Detailed neuropsychological testing covered seven cognitive domains: Visuospatial abilities (Benson Complex Figure Copy), Working memory (Craft Story 21 Immediate Recall), Delayed memory (Craft Story 21 Delayed Recall and Benson Complex Figure Recall), Language (Verbal Fluency–Animal), Attention/Processing speed (Trail Making Test–Part A), and Executive function (Trail Making Test–Part B).^24^ Z-scores for each test were computed using published age-, sex- and education-adjusted normative calculator;^24^ a global Z-score was then computed by averaging the Z-scores of all tests. Diagnoses of mild cognitive impairment (MCI) and dementia were made via consensus conference among three dementia specialists, using DSM-5 (Diagnostic and Statistical Manual of Mental Disorders–Fifth Edition) criteria^25^ for dementia, and modified Petersen criteria^26^ for MCI. Normal cognition was assigned when neither condition was present.

Subjective cognitive complaints (SCC) were assessed in both participants and informants using two validated approaches. First, a global SCC measure asked participants or informants: “Do you feel like your (or your family member’s) memory is becoming worse?” An affirmative response to this initial question prompted two follow-up questions to assess worry (’Does this worry you?’) and help-seeking intention (’Have you spoken to or intend to speak to a doctor about the memory concerns?’). This global measure of SCC has been validated in previous studies^27,28^ and shown to be useful for capturing early symptoms of cognitive decline.^29–33^

Second, the Everyday Cognition (ECog-12) questionnaire^34^ measured SCC across six cognitive domains, administered to both participants and informants. This 12-item instrument measures perceived changes in Memory, Language, Visuospatial Abilities, Planning, Organization, and Attention, with two items per domain. For each item, respondents compared the participant’s current ability to that of 10 years prior, allowing each participant to serve as their own baseline. Ratings were made on a 4-point scale: 1=better or no change, 2=occasionally worse, 3=consistently a little worse, 4=consistently much worse. For this analysis, responses were dichotomized to indicate the presence (a rating of 2–4) or absence (a rating of 1) of SCC in each domain. ECog-12 has been validated for its ability to capture a wide range of everyday cognitive symptoms^34^ and to reflect early symptoms of cognitive decline.^28^

### Statistical analyses

All statistical analyses were conducted in R (version 4.5.1). The analytical process involved two main stages: re-weighting the study sample to match Singapore’s older population (≥65 years), followed by descriptive and comparative analyses on the weighted data.

To provide population-representative estimates, we employed entropy balancing—a multivariate re-weighting technique—to calibrate the sample to national proportions for key demographic and clinical variables (age group, sex, ethnicity, education level, living arrangements, obesity, diabetes mellitus, hypertension, hyperlipidemia, dementia, as well as the known rate of undiagnosed dementia in Singapore). National statistics used for weighing are listed in Supplementary Material 1. The ebal R package was employed, configured with a constraint tolerance of 1e-14 and a maximum of 1,000,000 iterations to ensure strict convergence on population targets.

The weighting process generated a unique weight for each participant, which was then used to conduct subsequent weighted analyses. For Aim 1, we calculated the rate of previously undiagnosed CI, computed by the proportion of individuals with no self- or informant-reported history of a cognitive diagnosis among those who were diagnosed with CI (MCI or dementia) in our study. These rates, along with their 95% confidence intervals (95%CI), were calculated for CI overall and separately for MCI and dementia. For Aim 2, we examined the prevalence of SCC (both the global measure and the six domain-specific ECog-12 measures) across the diagnostic groups: Normal Cognition, MCI, and Dementia. All prevalence estimates are based on the weighted data for population-representative results. The survey and gtsummary packages in R were used to account for weighting in all calculations.

## RESULTS

### Sample Characteristics

The characteristics of the study sample, before and after weighting, are shown in Supplementary Material 2. After weighting, the final sample (n=1,856) precisely matched Singapore’s older population demographics (based on known national statistics as listed in Supplementary Material 1)—for example, the resulting profile was now 46.2% male, 82.9% of Chinese ethnicity, and 50.4% having below secondary education. Informant characteristics are detailed in Supplementary Material 3—the majority were spouses (42.4%) or children (35.7%), with 71.7% interacting daily with the participant.

### Prevalence and Undiagnosed Rate of Cognitive Impairment

After weighting, the population-representative prevalence of CI was 24.7% (95%CI 18.8–31.8). This total comprised a computed prevalence of 15.9% (95%CI 11.4–21.8) for MCI; and a calibrated point prevalence of 8.8% for dementia (weighted based on known national statistics as listed in Supplementary Material 1;^35^ weighted 95%CI 4.8–15.7).

Among participants with CI (n=458 in the weighted sample), 81.4% were previously undiagnosed (95%CI 65.6–90.9)—driven by a 97.9% undiagnosed rate for MCI (95%CI 94.1–99.3), while the rate for undiagnosed dementia was calibrated to 51.5% (weighted based on known national statistics as listed in Supplementary Material 1;^35^ weighted 95%CI 22.0–80.0). Detailed undiagnosed rates, before and after weighting, are presented in Supplementary Material 4.

### Subjective Cognitive Complaints and Help-Seeking Intention

Figure 1 shows the prevalence of SCC across diagnosis subgroups, with detailed results presented in Supplementary Material 5–6.

**Figure 1.**
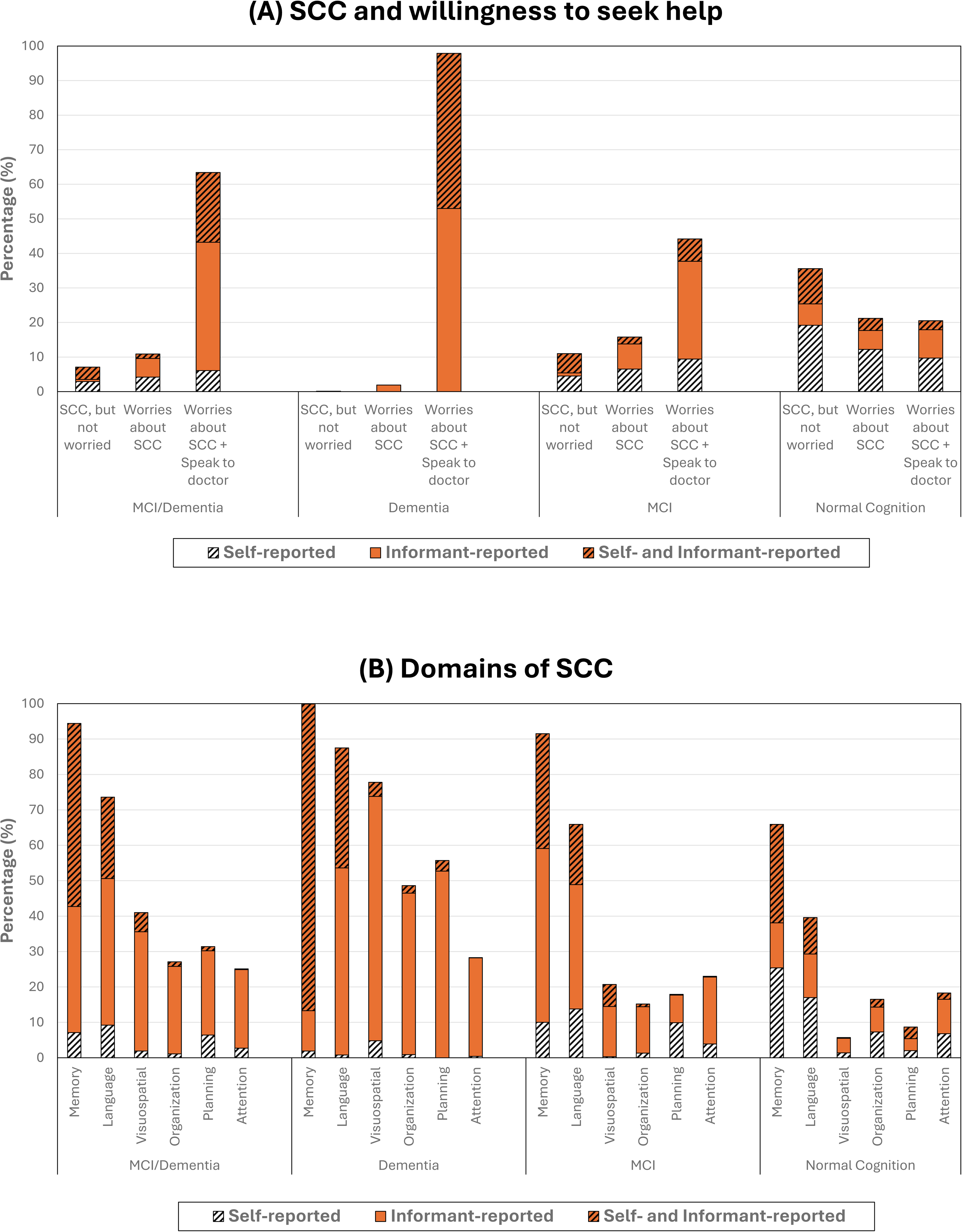
Proportion of older individuals with subjective cognitive concerns (SCC) as reported by self or informant, stratified by cognitive diagnosis (n=1,856 in the population-weighted sample). SCC, subjective cognitive concerns; MCI, mild cognitive impairment. *<u>Note</u>*: Figure (A) examines the proportion of individuals who were worried about their subjective cognitive symptoms. The third bar (i.e. Worries about SCC + Speak to doctor) reflects individuals who were worried about their subjective cognitive symptoms, and have spoken to or intended to speak to their doctors about their concerns. Figure (B) further breaks down the cognitive domains of SCC as reported by self or informant.

Figure 1A visualizes the prevalence of symptom awareness and help-seeking intention. Among all individuals with CI, 81.3% reported SCC (95%CI 63.6–91.5), and 63.3% expressed worry and intended to seek help (95%CI 47.5–76.7). In the dementia subgroup, virtually all reported SCC, with 97.9% also worried and intending to seek help (95%CI 87.2–99.7). For MCI, most reported SCC (70.9%, 95%CI 49.2–86.0), and 44.2% expressed worry and help-seeking intent (95%CI 27.1–62.8).

Figure 1B further visualizes the specific domains of SCC. Although Memory was the most frequently reported domain of concern among individuals with CI (94.4%, 95%CI 89.9–97.0), other non-memory domains were also noted, including Language (73.6%, 95%CI 29.2–84.2), Visuospatial abilities (41.0%, 95%CI 25.3–58.7), Planning (31.4%, 95%CI 17.1–50.4), Organization (27.1%, 95%CI 14.9–44.1), and Attention (24.9%, 95%CI 13.8–40.7).

As CI worsened, a notable pattern emerged in the source of SCC. As impairment progressed from MCI to dementia, informants became a more prominent source of SCC, particularly for non-memory domains (e.g. visuospatial abilities, organization, planning; Figure 1B) and help-seeking intentions (Figure 1A).

## DISCUSSION

### Summary of Findings

This community-representative study reveals a substantial burden of CI in Singapore, affecting nearly one in four older adults. Critically, over 80% of these cases were previously undiagnosed, with an alarmingly high rate for MCI (97.9%). Our central finding challenges the assumption that the diagnostic gap primarily stems from patient inaction^19^—over 80% of affected individuals had cognitive concerns, and nearly two-thirds were worried and intended to seek help. As impairment worsened, informants played an increasingly important role in reporting non-memory symptoms (e.g. planning, visuospatial abilities) and initiating help-seeking. Taken together, these findings highlight a stark discrepancy (high patient and informant awareness versus low diagnostic rates), raising a critical question: What other non-patient factors^21,22^ hinder these help-seeking individuals from obtaining a formal diagnosis?

### Interpretation of Findings

To our knowledge, this is one of the first community-representative studies to quantify the undiagnosed rates of CI, incorporating both MCI and dementia. Although the rate of undiagnosed dementia has been widely studied, literature on undiagnosed MCI has been scarcer. While alarming, our finding on the high rate of undiagnosed MCI (97.9%) is remarkably consistent with a recent US study, where over 90% of expected MCI cases in the community were reported to be undiagnosed.^3^ This consistency lends external validity to our estimates and underscores a global challenge in detecting early CI. Crucially, this diagnostic gap does not appear to stem from the condition being silent or benign. On the contrary, we found that a sizeable proportion of individuals with MCI (44.2%) were worried and intended to seek help, which challenges the common misconception that MCI is trivial—a belief perhaps perpetuated by the misnomer ‘mild’ itself. In fact, our findings are consistent with extant literature demonstrating that MCI is associated with significant psychological distress, challenges in complex daily activities, and substantial caregiver burden.^36,37^ As the CI worsens, our findings also highlighted the increasingly prominent role of informants especially for reporting non-memory symptoms (e.g. planning, visuospatial abilities) and for initiating the process of seeking help. This shift is a classic manifestation of anosognosia, where individuals increasingly lose awareness of their own deficits as the CI progresses,^38^ necessitating that informants become the primary agents to recognize these deficits and seek help on the individuals’ behalf.

A striking finding of this study is the profound disconnect—over 80% of CI cases were undiagnosed, even though nearly two-thirds of these individuals were worried and intended to consult a doctor. This challenges the prevailing narrative that attributes the diagnostic gap primarily to patient-level factors such as denial or poor awareness.^19^ While these patient factors remain relevant in some cases, our data suggest they may no longer be the principal drivers of the diagnostic gap in Singapore. Instead, the high symptom awareness and willingness to seek help likely reflect the positive impact of Singapore’s sustained public health initiatives. Following a landmark finding of 10% dementia prevalence in 2013,^39^ Singapore undertook concerted, decade-long governmental efforts to improve public dementia literacy, notably through the national campaign of “Dementia Friendly Singapore”.^40^ These efforts appear to have empowered the public to recognize and act on cognitive symptoms, as reflected by a follow-up study showing the proportion of undiagnosed dementia cases fell significantly from 70.6% in 2013 to 51.5% in 2023.^35^ This success suggests that the next frontier for improving diagnostic rates may lie not in generating further demand, but in enhancing the system’s capacity^21^ to absorb this rising tide of help-seekers.

### Policy Implications

Essentially, our findings point to an opportunity for a strategic evolution in public health policy, building upon past success to now focus on strengthening the healthcare system’s capacity to respond.^21^ A cornerstone of this strategy could be the empowerment of primary care and the clarification of the diagnostic pathway. Currently, systemic obstacles can pose challenges, creating a potential risk where motivated individuals may not complete their journey to diagnosis.^21,22^ For instance, primary care physicians may face challenges such as time constraints and a lack of routine cognitive assessment tools, while the clinical pathway from an initial concern to a specialist diagnosis can present complexities.^22^ Policies can help address these challenges. To mitigate time constraints, brief cognitive screening may be implemented for high-risk groups in primary care, perhaps by embedding validated tools into routine workflows.^23,41^ To address pathway complexity, strengthening and formalizing integrated care pathways is a key next step. This may entail further refining national-level guidelines to clearly define the roles of primary care, community services, and specialist memory clinics, ensuring that referrals are seamlessly managed, supported by shared patient records, and followed-up with standardized protocols.^42^

Another system-level opportunity is to formally empower informants as essential partners in detection. As our findings show, the informant’s role becomes paramount as CI worsens, yet this vital resource can be underutilized due to a fundamental communication gap. Older adults often attend appointments alone, physically separating the concerned informant from the clinician. This creates a critical risk: a clinical encounter that relies solely on a patient’s self-report may miss crucial information, leading to false reassurance and a delayed diagnosis. Policy can bridge this communication gap by creating formal channels for informant contact,^43^ such as secure messaging portals for informants to raise concerns, or protocols for clinicians to proactively engage designated family members (e.g. pre-visit questionnaires). By systematically creating a ‘virtual door’ for informants, the system can capture crucial diagnostic information that might otherwise be missed.

However, creating this channel is only half the solution. It is also crucial to equip informants with the confidence and specific language to articulate their concerns effectively,^44^ ensuring their observations are not dismissed as vague worries or misinterpreted as “normal aging”. To this end, public health campaigns can evolve beyond the general message of “see a doctor for memory loss” to co-target family members, educating them on the broader spectrum of cognitive symptoms to report to clinicians—for instance, by providing concrete examples of changes in organization (e.g. trouble managing finances), attention (e.g. losing the thread of conversations), and visuospatial abilities (e.g. difficulty navigating familiar places). Ultimately, this strategy empowers informants to articulate their observations with clarity, transforming their input into valuable clinical data that can enrich the diagnostic conversation.

### Global Implications and Generalizability

While our specific finding of high patient awareness is likely influenced by Singapore’s unique and successful public health context, the underlying principle is highly generalizable. Our study illustrates a ‘paradox of success’, which we conceptualize as a dynamic two-stage model as illustrated in Figure 2. In Stage 1 (Awareness Gap), the primary bottleneck for diagnosis is at the patient and community level due to factors like low awareness or stigma. As public health initiatives succeed in empowering patients, this barrier is overcome, leading to Stage 2 (Capacity Gap), where the bottleneck shifts to systemic capacity. This provides a crucial lesson for other nations, showing that policy must evolve in step with public readiness; as awareness campaigns succeed, the focus must naturally shift from generating demand to strengthening the healthcare system’s capacity to absorb it.

**Figure 2.**
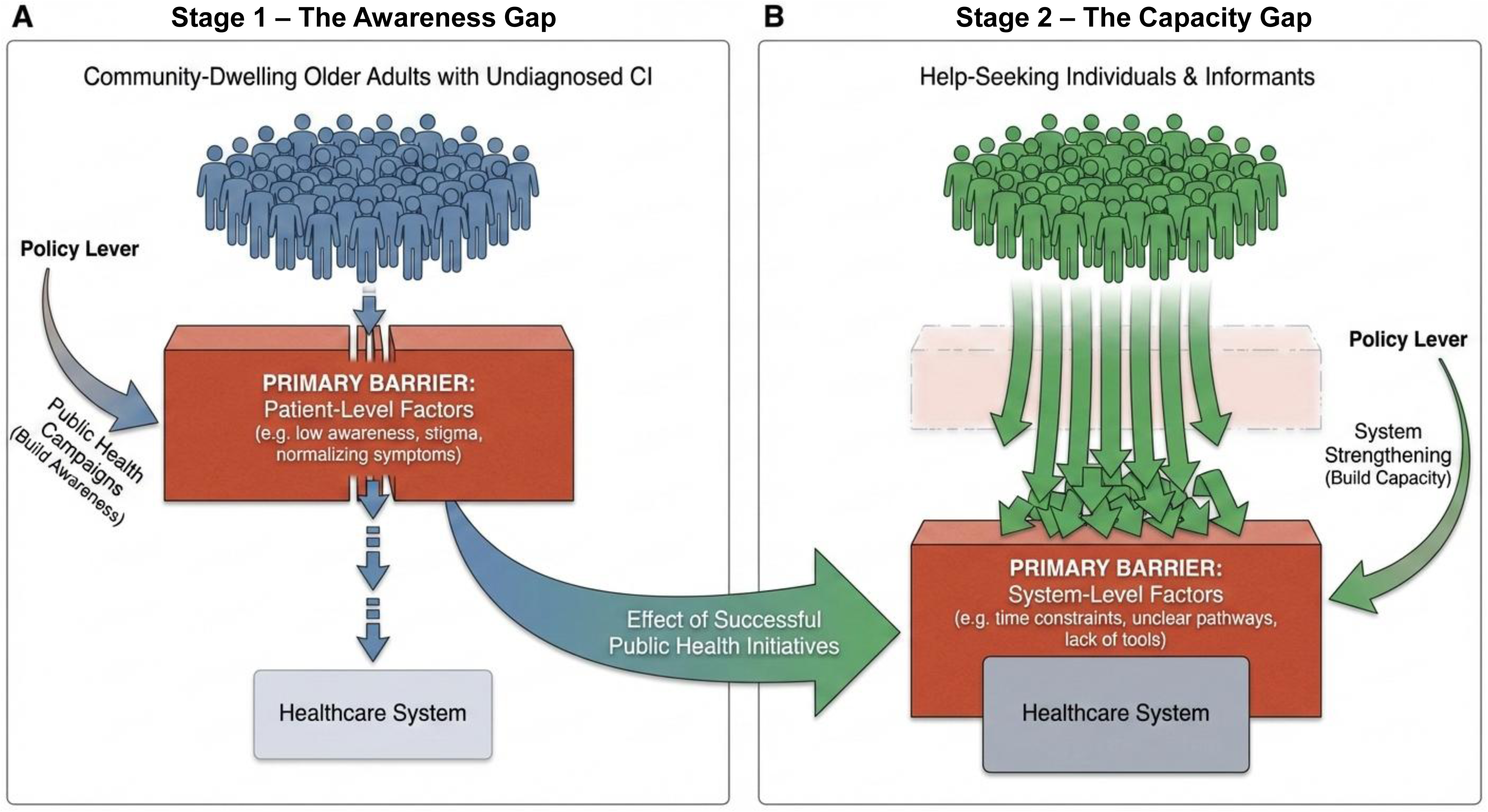
A dynamic model of shifting barriers in the diagnosis of cognitive impairment CI, cognitive impairment. *<u>Note:</u>* Image generated using Google Gemini 3.0 Pro (Nano Banana Pro) on 21st December 2025 based on user-provided prompts. The final output was reviewed and verified for accuracy by the authors.

Furthermore, our methodology provides a Diagnostic Landscape Assessment blueprint for other health systems. This blueprint involves three key steps: (1) Quantify the true diagnostic gap using rigorous, community-representative sampling, not clinic-based data; (2) Simultaneously measure help-seeking readiness, assessing both symptom awareness and the intention to consult a clinician; and (3) Triangulate perspectives by collecting this information from both the individual and a knowledgeable informant. By applying this framework, another health system can accurately identify its primary barrier—whether it is a patient awareness gap or a systemic capacity gap—and thereby tailor its public health policies to the most pressing local need.

### Limitations

Several limitations are notable. First, the research participants can arguably differ from those who did not participate. We mitigated this limitation by employing diverse sources of community recruitment to engage a broader cross-section of the community (e.g. by actively reaching out to community-dwelling individuals through our networks of community partners and our 14 community roadshows), rather than relying solely on conventional research invitations that might disproportionately attract health-seeking individuals. Second, the dataset included only community-dwelling individuals (i.e. excluded those already in nursing homes), which may reduce its representativeness; however, this risk is low as only a small minority of older adults in Singapore (2.7%) reside in nursing homes.^45^ Third, the small proportion of non-Chinese participants constraining our re-weighting process to a Chinese versus non-Chinese variable, potentially obscuring distinct patterns in minority groups (e.g. Malay, Indian, Eurasian). Fourth, the number of dementia cases was small, resulting in wide confidence intervals for this subgroup’s estimates.

### Conclusions

This study reveals a high rate of undiagnosed cognitive impairment in Singapore, despite high symptom awareness and help-seeking intent. The findings suggest that successful public health campaigns have shifted the bottleneck from community awareness to healthcare system capacity. This presents an opportunity for a strategic policy shift to meet the rising demand for diagnosis—by empowering primary care with efficient case-finding tools, formalizing integrated diagnostic pathways, and establishing channels for family informants’ involvement. Based on the findings, we conceptualized a ‘paradox of success’ model, providing a framework for other health systems to assess their position and adapt policy as public engagement grows.

## Data Availability

The data used in this study contains sensitive information about the study participants and they did not provide consent for public data sharing. The current approval by the SingHealth Centralized IRB of Singapore (reference number: 2021/2590) does not include data sharing.

## ACKNOWLEDGEMENTS

We thank the following persons for their assistance in recruiting study participants: Yanling Tan, Alcantara Leicester Shawn, Kai Xin Choo, Megan Cheng Mun Choy, Xin Tong Tan, Lydia Jia En Cheong, Jane Mee Chin Liew, Xiao Hui Ng, and Spencer Peng Ming Yuen. We also thank the following community organizations for their support in participant recruitment: Thye Hua Kwan Moral Charities (AMK 645, Bukit Merah View, Beo Crescent), Kreta Ayer Senior Activity Centres, NTUC Health Active Ageing Centre (Lengkok Bahru), Agency for Integrated Care (Caregiving & Community Mental Health Division), Silver Generation Office, People’s Association, Precious Active Ageing Centre, Montfort Care, Yong-en Care Centre, Presbyterian Community Services, Lions Befrienders Service Association Singapore.

## ETHICAL STANDARDS

The study received ethical approval from the SingHealth Centralized IRB of Singapore (reference number: 2021/2590). Informed consent was obtained from all participants. Before obtaining informed consent, the mental capacity of participants was briefly assessed in accordance with the Mental Capacity Act of Singapore. In the event there were concerns about mental capacity, informed consent by proxy was then obtained from legally authorized next-of-kin.

## DECLARATION OF GENERATIVE AI AND AI-ASSISTED TECHNOLOGIES IN THE WRITING PROCESS

Figure 2 was generated using Google Gemini 3.0 Pro (Nano Banana Pro) on 21st December 2025 based on user-provided prompts. The final output was reviewed and verified for accuracy by the authors.

