## Supplementary Material for "Undiagnosed cognitive impairment and willingness to seek help: Community-representative study from Singapore"

**Supplementary Material 1.** National statistics related to older adults ≥65 years in Singapore.

| Characteristic | National statistics | Reference |
|---|---|---|
| Age group in years | 65–69 years: 35.3%<br>70–74 years: 27.4%<br>75–79 years: 17.9%<br>80–84 years: 10.3%<br>85+ years: 9.2% | Singapore Department of Statistics [1] |
| Sex | Male: 46.2%<br>Female: 53.8% | Singapore Department of Statistics [1] |
| Ethnicity | Chinese: 82.9%<br>Non-Chinese: 17.1%s | Singapore Department of Statistics [1] |
| Education status | Below secondary: 50.4%<br>Secondary: 21.6%<br>Post secondary (non-tertiary): 11.9%<br>Diploma and Professional qualification: 7.4%<br>University: 8.7% | Singapore Department of Statistics [2] |
| Living alone | 11.4% | Singapore Department of Statistics [3] |
| Obesity (BMI≥30 kg/m$^2$) | 9.6% | National Population Health Survey 2024 [4] |
| Diabetes mellitus | 22.3% [a] | National Population Health Survey 2023 [5] |
| Hypertension | 47.7% [a] | National Population Health Survey 2023 [5] |
| Hyperlipidemia | 50.9% [a] | National Population Health Survey 2023 [5] |
| Dementia | 8.8% | TheWell-being of the Singapore Elderly (WiSE) 2023 [6] |
| Rate of undiagnosed dementia | 51.5% | TheWell-being of the Singapore Elderly (WiSE) 2023 [6] |

[a] National statistics based on self-reports were used, to align with the self-report disease status in the current dataset.

**Supplementary Material 2.** Comparison of sample characteristics before and after weighting (n=1,856)

| Characteristic | Unweighted Sample (n=1,856) | Weighted Sample (n=1,856) |
|---|---|---|
| Age group in years, n (%) | | |
| 65–69 | 646 (34.8) | 655 (35.3) [1] |
| 70–74 | 649 (35.0) | 508 (27.4) [1] |
| 75–79 | 351 (18.9) | 331 (17.9) [1] |
| 80–84 | 157 (8.5) | 191 (10.3) [1] |
| 85+ | 53 (2.9) | 170 (9.2) [1] |
| Education status, n (%) | | |
| Below secondary | 325 (17.5) | 936 (50.4) [1] |
| Secondary | 738 (39.8) | 400 (21.6) [1] |
| Post secondary (non-tertiary) | 296 (15.9) | 221 (11.9) [1] |
| Diploma and Professional qualification | 189 (10.2) | 138 (7.4) [1] |
| University | 308 (16.6) | 162 (8.7) [1] |
| Male sex, n (%) | 688 (37.1) | 858 (46.2) [1] |
| Chinese ethnicity, n (%) | 1,735 (93.5) | 1,539 (82.9) [1] |
| Living alone, n (%) | 285 (15.4) | 211 (11.4) [1] |
| Obesity, n (%) | 107 (5.8) | 178 (9.6) [1] |
| Known diabetes mellitus, n (%) | 564 (30.4) | 414 (22.3) [1] |
| Known hypertension, n (%) | 1,170 (63.0) | 885 (47.7) [1] |
| Known hyperlipidemia, n (%) | 1,551 (83.6) | 945 (50.9) [1] |
| Known heart disease, n (%) | 387 (20.9) | 261 (14.1) |
| Known neurological condition, n (%) | 154 (8.3) | 212 (11.4) |
| Known psychiatric condition, n (%) | 112 (6.0) | 120 (6.4) |
| NTB global Z-score, mean (SD) | -0.3 (0.6) | -0.5 (0.8) |
| Presence of cognitive impairment, n (%) [95%CI] | 255 (13.7) [12.2–15.4] | 458 (24.7) [18.8–31.8] |
| Breakdown of cognitive diagnosis, n (%) [95%CI] | | |
| Normal cognition | 1,601 (86.3) [84.6–87.8] | 1,398 (75.3) [68.2–81.2] |
| Mild cognitive impairment | 207 (11.2) [9.8–12.7] | 295 (15.9) [11.4–21.8] |
| Dementia | 48 (2.6) [1.9–3.4] | 163 (8.8) [4.8–15.7] [1] |

SD, standard deviation; NTB, neuropsychological test battery; 95%CI, 95% confidence interval.

[1] To provide population-representative estimates, our sample's distribution was weighted to match known national statistics among Singapore's older population (⩾65 years). The national statistics used in this weighting procedure are further presented in Supplementary Material 1.

**Supplementary Material 3.** Characteristics of the study informants (n=1,856)

| Variable | Unweighted Sample (n=1,856) | Weighted Sample (n=1,856) |
|---|---|---|
| Age, mean (SD) | 62.4 (14.0) | 60.2 (14.1) |
| Male sex, n (%) | 655 (35.3) | 588 (31.7) |
| Relationship to the participant, n (%) | | |
| Spouse | 897 (48.3) | 786 (42.4) |
| Children/Children-in-law | 506 (27.3) | 663 (35.7) |
| Other family member or relative | 255 (13.7) | 240 (12.9) |
| Friend/Neighbour | 167 (9.0) | 134 (7.2) |
| Care worker/Domestic helper | 31 (1.7) | 33 (1.8) |
| Frequency of contact with participant, n (%) | | |
| Daily | 1,340 (72.2) | 1,330 (71.7) |
| At least once a week | 405 (21.8) | 404 (21.8) |
| At least once a month | 89 (4.8) | 91 (4.9) |
| Once every few months or lesser | 22 (1.2) | 29 (1.6) |

SD, standard deviation.

**Supplementary Material 4.** Detailed results on the undiagnosed rates of cognitive impairment

| History of cognitive impairment | Unweighted Sample | Weighted Sample |
|---|---|---|
| Subset: MCI/Dementia | | |
| No known history of cognitive impairment, n (%) [95%CI] | 224 (87.8) [83.0–91.5] | 373 (81.4) [65.5–91.0] |
| Known history of cognitive impairment, n (%) [95%CI] | 31 (12.2) [8.5–17.0] | 85 (18.6) [9.0–34.5] |
| Subset: MCI | | |
| No known history of cognitive impairment, n (%) [95%CI] | 196 (94.7) [90.4–97.2] | 289 (97.9) [94.1–99.3] |
| Known history of cognitive impairment, n (%) [95%CI] | 11 (5.3) [2.8–9.6] | 6 (2.1) [0.7–5.9] |
| Subset: Dementia | | |
| No known history of cognitive impairment, n (%) [95%CI] | 28 (58.3) [43.3–72.1] | 84 (51.5) [22.0–80.0] |
| Known history of cognitive impairment, n (%) [95%CI] | 20 (41.7) [27.9–56.7] | 79 (48.5) [20.0–78.0] |

95%CI, 95% confidence interval.

**Supplementary Material 5.** Weighted results on the types of subjective cognitive concerns as reported by self or informant, stratified by cognitive diagnosis.

| Source of SCC [1] | Type of SCC | | |
|---|---|---|---|
| | SCC, but not worried | Worries about SCC | Worries about SCC + Speak to doctor |
| MCI/Dementia (n=458) | | | |
| self | 13 (2.9) [1.5–5.7] | 19 (4.2) [2.0–8.6] | 28 (6.1) [1.6–20.1] |
| informant | 3 (0.6) [0.2–1.9] | 25 (5.4) [2.3–12.3] | 170 (37.1) [23.1–53.6] |
| self and informant | 16 (3.6) [1.8–7.0] | 6 (1.3) [0.3–5.8] | 92 (20.2) [8.2–41.5] |
| **SUBTOTAL [2]** | **32 (7.1) [4.2–11.6]** | **50 (10.9) [6.0–18.8]** | **290 (63.3) [47.5–76.7]** |
| Dementia (n=163) | | | |
| self | 0 (0.1) [0.0–0.1] | 0 (0.0) [0.0–0.0] | 0 (0.0) [0.0–0.0] |
| informant | 0 (0.0) [0.0–0.0] | 3 (1.9) [0.2–13.4] | 87 (53.0) [22.8–81.2] |
| self and informant | 0 (0.0) [0.0–0.0] | 0 (0.0) [0.0–0.3] | 73 (44.9) [17.1–76.4] |
| **SUBTOTAL [2]** | **0 (0.1) [0.0–0.1]** | **3 (2.0) [0.3–13.3]** | **160 (97.9) [87.2–99.7]** |
| MCI (n=295) | | | |
| Self | 13 (4.5) [2.2–8.9] | 19 (6.5) [3.0–13.3] | 28 (9.4) [2.6–29.0] |
| Informant | 3 (0.9) [0.3–2.9] | 22 (7.3) [2.9–17.5] | 84 (28.3) [13.8–49.3] |
| self and informant | 16 (5.6) [2.8–10.8] | 6 (2.0) [0.4–9.0] | 19 (6.5) [3.2–12.6] |
| **SUBTOTAL [2]** | **32 (10.9) [6.4–18.0]** | **47 (15.8) [8.6–27.2]** | **131 (44.2) [27.1–62.8]** |
| Normal Cognition (n=1398) | | | |
| self | 268 (19.2) [15.5–23.4] | 171 (12.2) [9.6–15.4] | 136 (9.7) [7.2–12.9] |
| informant | 87 (6.2) [4.0–9.6] | 76 (5.5) [3.6–8.1] | 115 (8.2) [4.6–14.1] |
| self and informant | 142 (10.2) [6.8–15.1] | 49 (3.5) [2.2–5.5] | 37 (2.6) [0.9–7.3] |
| **SUBTOTAL [2]** | **497 (35.6) [30.4–41.2]** | **296 (21.2) [17.6–25.3]** | **287 (20.5) [15.6–26.5]** |

[1] Values in the table indicate n (%) [95% Confidence Interval], with the percentages computed based on the total number within each diagnosis subset.
[2] The remaining numbers (making up to 100%) reflect individuals who did not report SCC within the diagnosis subset.

**Supplementary Material 6.** Weighted results on the domains of subjective cognitive concerns as reported by self or informant, stratified by cognitive diagnosis.

| Source of SCC [1] | Domain of SCC | | | | | |
|---|---|---|---|---|---|---|
| | Memory | Language | Visuospatial | Organization | Planning | Attention |
| MCI/Dementia (n=458) | | | | | | |
| self | 33 (7.1) [3.6–13.6] | 42 (9.2) [5.0–16.3] | 9 (1.9) [0.4–8.3] | 5 (1.1) [0.5–2.5] | 29 (6.4) [1.7–21.0] | 12 (2.7) [13–5.5] |
| informant | 163 (35.6) [21.4–53.0] | 190 (41.4) [25.7–59.0] | 154 (33.7) [18.8–52.7] | 113 (24.7) [12.9–42.1] | 109 (23.8) [11.0–44.3] | 101 (22.1) [11.5–38.3] |
| self and informant | 237 (51.7) [35.6–67.5] | 106 (23.0) [11.9–39.9] | 25 (5.4) [1.2–20.9] | 6 (1.3) [0.4–3.9] | 6 (1.2) [0.3–5.3] | 1 (0.2) [0.0–0.7] |
| **SUBTOTAL [2]** | **433 (94.4) [89.9–97.0]** | **337 (73.6) [59.2–84.2]** | **188 (41.0) [25.3–58.7]** | **124 (27.1) [14.9–44.1]** | **144 (31.4) [17.1–50.4]** | **114 (24.9) [13.8–40.7]** |
| Dementia (n=163) | | | | | | |
| self | 3 (1.9) [0.2–13.4] | 1 (0.8) [0.1–5.6] | 8 (4.8) [0.8–23.4] | 1 (0.9) [0.2–3.7] | 0 (0.0) [0.0–0.0] | 1 (0.4) [01–3.3] |
| informant | 19 (11.4) [4.0–28.0] | 86 (52.8) [23.6–80.2] | 113 (69.0) [39.3–88.4] | 75 (45.6) [19.0–75.1] | 86 (52.7) [23.9–79.9] | 45 (27.8) [9.8–57.8] |
| self and informant | 141 (86.6) [69.0–94.9] | 55 (33.9) [11.7–66.5] | 6 (4.0) [0.9–15.8] | 3 (2.1) [0.5–8.7] | 5 (3.0) [0.5–.5] | 0 (0.1) [0.0–0.5] |
| **SUBTOTAL [2]** | **163 (99.9)** [99.0–100.0] | **143 (87.5)** [64.0–96.5] | **127 (77.7)** [46.5–93.3] | **79 (48.6)** [20.7–77.5] | **91 (55.7)** [26.6–81.4] | **46 (28.3)** [10.1–58.2] |
| MCI (n=295) | | | | | | |
| Self | 29 (10.0) [4.9–19.4] | 41 (13.8) [7.3–24.5] | 1 (0.3) [0.1–0.9] | 4 (1.3) [0.5–3.3] | 29 (9.9) [2.7–30.3] | 11 (3.9) [1.8–8.3] |
| informant | 145 (49.1) [31.3–67.1] | 104 (35.1) [18.6–56.1] | 42 (14.2) [4.4–36.9] | 39 (13.1) [3.8–36.7] | 23 (7.8) [2.8–19.8] | 56 (18.9) [7.7–39.7] |
| self and informant | 96 (32.4) [19.1–49.2] | 50 (17.0) [7.2–34.8] | 18 (6.2) [0.9–32.8] | 2 (0.8) [0.1–5.2] | 1 (0.2) [0.0–1.5] | 1 (0.2) [0.1–1.1] |
| **SUBTOTAL [2]** | **270 (91.4) [84.4–95.5]** | **194 (65.9) [48.0–80.1]** | **61 (20.7) [8.1–43.5]** | **45 (15.2) [5.2–37.0]** | **53 (18.0) [7.9–35.9]** | **68 (23.1) [10.9–42.4]** |
| Normal Cognition (n=1398) | | | | | | |
| Self | 355 (25.4) [21.0–30.4] | 237 (17.0) [13.6–21.0] | 20 (1.4) [0.9–2.2] | 102 (7.3) [4.1–12.6] | 28 (2.0) [1.2–3.4] | 96 (6.8) [5.0–9.4] |
| informant | 177 (12.7) [9.5–16.7] | 172 (12.3) [8.5–17.5] | 57 (4.1) [1.4–11.2] | 97 (7.0) [3.4–13.8] | 48 (3.4) [2.0–5.8] | 135 (9.7) [5.8–15.6] |
| self and informant | 389 (27.8) [23.0–33.2] | 144 (10.3) [6.3–16.4] | 3 (0.2) [0.1–0.6] | 30 (2.2) [1.0–4.5] | 46 (3.3) [0.9–11.8] | 25 (1.8) [0.7–4.1] |
| **SUBTOTAL [2]** | **922 (65.9) [60.2–71.3]** | **553 (39.6) [33.9–45.5]** | **79 (5.7) [2.6–11.7]** | **230 (16.4) [11.3–23.3]** | **122 (8.7) [5.1–14.6]** | **255 (18.3) [13.7–23.9]** |

[1] Values in the table indicate n (%) [95% Confidence Interval].
[2] The remaining numbers (making up to 100%) reflect individuals who did not report SCC in each domain.
